# EEG signatures at ascending vs descending propofol vs sevoflurane concentrations analyzed with BIS and a State-Chart Score

**DOI:** 10.64898/2026.10.03.26364648

**Authors:** C. Sun, P.O. Michel, A. Rigouzzo, J. Loh, D. Longrois, I. Constant, D. Holcman

**Affiliations:** Group of Data Modeling, Computational Biology and Predictive Medicine, Institut de Biologie (IBENS); École Normale Supérieure; Université PSL Paris, France; UPEC, Creteil France. SignalMedPlus, 229 rue St Honoré, 75001 Paris France; Département d’Anesthésie, Hôpital Armand Trousseau, Assistance Publique - Hôpitaux de Paris, Paris, France; Department of Anesthesia and Intensive Care, DMU (Département Médico-Universitaire) PARABOL, Assistance Publique-Hôpitaux de Paris, GHU (Groupe Hospitalo-Universitaire) Nord, Hôpital Louis Mourier, Université Paris Cité, INSERM 1148, France; Signalmed+, 229 rue St Honoré, 75001 Paris France

**Keywords:** General anesthesia, EEG, Statistical analysis, Alpha rhythms, Iso-electric suppressions, State-Chart, Propofol, Sevoflurane

## Abstract

**Background:** Depth of anesthesia is routinely monitored using electroencephalographic (EEG) indices such as the Bispectral Index (BIS), which reduce cortical activity to a single numerical real-time series. However, these scalar descriptors fail to capture inter-individual variability, drug-specific EEG signatures, and transitions between brain states.

**Methods:** We analyzed transient EEG patterns during general anesthesia induced by propofol and sevoflurane in 89 patients. Iso-electric suppressions (IES), *α*-suppressions, and spectral power in the *α* and *δ* bands were quantified across controlled concentration plateaus. We compared conventional BIS monitoring with a novel State-Chart representation mapping EEG activity into discrete depth states.

**Results:** Increasing hypnotic concentration produced a consistent shift from *α*-dominant to *δ*-dominant activity and a reduction in maximal *α* frequency. IES occurred predominantly at high propofol concentrations and remained infrequent under sevoflurane. Compared with BIS, the State-Chart showed a marked reduction in temporal variability (0.5 ± 0.6 vs 4.0 ± 2.9; *p* < 10^−90^). Importantly, our State-Chart separates stable from transition regimes and discriminates identical concentration levels. Statistics computed over prior dosing (ascending vs descending), reveals refined clustering not captured by BIS.

**Conclusions:** Anesthesia depth drugs concentrations vs EEG effects is more accurately described by structured EEG state transitions than by a single numerical index. The State-Chart provides a stable, physiologically interpretable representation of brain activity and reveals transition states preceding deep cortical suppression, supporting individualized and predictive monitoring of anesthesia.

## 1 Introduction

Documenting and avoiding both under- and over-dosage of hypnotic drugs remains a challenge in modern anesthetic practice. In routine clinical settings, anesthetic depth is adjusted empirically based on pharmacological targets (e.g., EC50/EC95), hemodynamic tolerance, and responsiveness to nociceptive stimuli. Hypnosis is commonly achieved using agents such as propofol and sevoflurane [1, 2, 3]. Although both are GABA-ergic anesthetics, they differ in their mechanisms of action: propofol primarily enhances inhibitory synaptic transmission via GABA_*A*_ receptor modulation, whereas sevoflurane additionally affects glycine receptors and potassium channels, leading to broader modulation of neuronal excitability [4]. These mechanistic differences result in distinct cortical electroencephalographic (EEG) signatures, complicating the interpretation of brain monitoring [5, 3].

Electroencephalography provides a direct, non-invasive measure of cortical activity and reveals spectral (with the spectrogram) changes during anesthesia, including transitions from *α* oscillations to *δ*-dominant activity as sedation deepens [6]. To simplify this complexity, processed EEG monitors such as the Bispectral Index (BIS) and Patient State Index (PSI) reduce brain activity to a single numerical value in time. These indices were statistically calibrated to clinical endpoints (e.g., loss of responsiveness or probability of recall), leading to widely adopted target ranges assumed to represent adequate anesthesia across different agents. Indeed, these statistical associations, coupled with a statistical approach of pharmacology (e.g. ED50/ED90 for a given drug) resulted in description of an adequate anesthetic state for values between 45-60 for the BIS and 25-50 for the PSI, independently of the GABA-ergic drug used.

However, this scalar representation has important limitations. First, it does not adequately account for inter-individual variability in brain sensitivity to anesthetics. Second, the relationship between drug concentration and EEG activity is highly non-linear. Third, different anesthetic agents with distinct neurophysiological effects often produce similar BIS values, preventing identification of drug-specific signatures. Finally, the use of fixed thresholds implicitly assumes stationary brain states and does not capture transitions between anesthetic regimes, such as the progression toward cortical suppression. Experimental and clinical observations suggest that anesthesia is better characterized as a sequence of transitions between oscillatory brain states rather than a continuous decrease in brain activity. Transient EEG patterns, including *α*-suppressions, iso-electric suppressions (IES), and spectral shifts, provide direct information about cortical responsiveness and brain dynamics [7, 6]. Characterizing these patterns across anesthetic concentrations may therefore improve the understanding of heterogeneous patient responses [8].

The objective of this study was twofold. First, we quantified EEG signatures associated with ascending and descending concentrations of propofol and sevoflurane in children and adults, focusing on spectral power distribution in the *α* and *δ* bands and the occurrence of *α*- and isoelectric suppressions. Second, we compared conventional BIS monitoring with a novel discrete State-Chart representation that maps EEG activity into ordered brain states. We hypothesized that (i) propofol and sevoflurane induce distinct EEG state transitions despite similar clinical depth targets, and (ii) a state-based representation provides a more stable and physiologically interpretable characterization of anesthesia than a single numerical index, particularly for identifying transition regimes and variability in brain response.

## 2 Materials and Methods

### 2.1 Statement about General Anesthesia protocol

This prospective observational randomized study included children and young adults patients receiving a scheduled elective procedure requiring general anesthesia under sevoflurane and propofol. We used a similar cohort as the one described in [9]. For each patient, two or more plateaus of 10 minutes duration were applied with a constant concentration of hypnotic. This study was approved by the Ethics Committee CPP Saint Antoine, Paris, France (Approval number: 04605).

Written and informed consent was obtained from children and their parents. The protocol has been therefore performed in accordance with the ethical standards laid down in 1964 declaration of Helsinki and its later amendments. A group of 89 patients ranging from 6 to 38 years old, ASA 1 or 2, was used in this study. Data were collected between 2005 and 2006. The trial was later on registered and completed on September 2016 (ClinicalTrials.gov ID: NCT02893904). We used MATLAB R2021a software to perform the statistical analysis. The significance level used in this study was *α*=0.05. The values were expressed in mean ± standard deviation.

### 2.2 General anesthesia protocol

All subjects received premedication with oral hydroxyzine (1 mg.*kg*^−1^) before surgery. During induction, propofol was injected with an initial target concentration of 6 *µg*.*ml*^−1^. For sevoflurane, induction was performed at 6% with 100% oxygen. Both induction regimens were followed by continuous administration of remifentanil (0.25 *µ*g.*kg*^−1^.*min*^−1^). The patients were then intubated with cuffed tracheal tube after a single dose atracurium besilate (0.5 mg.*kg*^−1^) and mechanically ventilated with an air-oxygen mixture and a breathing frequency ranging from 14 to 20 *min*^−1^ to obtain an *ET*_*CO*2_ between 30 and 35 mmHg.

The propofol concentration was kept between 2 and 6 *µg*.*ml*^−1^ according to the Schnider model for post-pubertal subjects and Kataria model for pre-pubertal subjects [10]. End-tidal concentrations of sevoflurane were maintained between 1 and 5%.

The decision to administer extra medication was left to the discretion of the anesthesiologist in charge following the institution standard of care. The frontal cortical EEG monitoring was performed by Bispectral index (BIS), monitor (Medtronic). Other monitored variables were, pulse oxygen saturation (SpO_2_), heart rate (CF), systolic and diastolic blood pressures (SBP, DBP), mean arterial pressure (MAP), and oesophageal temperature.

### 2.3 EEG recordings and pre-processing

EEG were recorded using the Brain-Quick program stem II (Micromed, France) with a single channel record from electrodes placed on the forehead, left and right (reference) mastoids [11]. We segmented the mechanical noise using the EEG signal power within a sliding window of length 10 seconds and 50% overlap. A time-segment *P*_*i*_ was considered artifactual when the spectral power within the window exceeded three times the median absolute value,

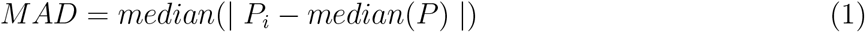

for *i* = 1, 2, .., *N*, where *N* is the number of windows. Segmented artifacts are then corrected using the classical Wavelet Quantile Normalization method [12, 13].

### 2.4 IES and *α*S segmentation

Iso-electric suppressions (IES) and *α*− suppressions (*α*S) were segmented following a procedure described in [7]: we first filtered the EEG signal *S*(*t*) in the range 8−16 Hz leading to the signal *S*_*α*_(*t*). We normalized *S*_*α*_(*t*) by its Root-Mean-Square leading to *Ŝ*_*α*_(*t*). We computed the difference *D*(*t*) and *D*_*α*_(*t*) between the respective upper and lower envelops local maxima and minima interpolation, defining two threshold values

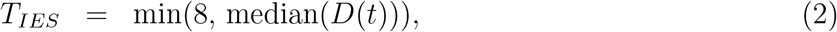

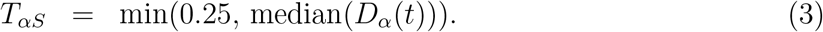

An EEG segment is classified as iso-electric (resp. *α*) suppression, when the amplitude of *Ŝ*_*α*_ is smaller than *T*_*αS*_, the amplitude of *S*(*t*) is smaller (resp. greater) than *T*_*IES*_ while the duration should exceed at least 1 second (resp. 0.5 second).

### 2.5 Mathematical indicators for the *α* and *δ* bands

- The *α*−band relative power *P*_*α*_(*t*) describes the power proportion of the concentration plateau in the [8, 12] Hz range with respect to [0.1, 45] Hz, as defined by the expression

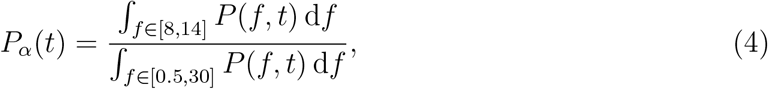

where *P* is the single channel EEG spectrogram obtained from a Fourier transform on a 40 seconds sliding window and 75% overlap. To obtain the mean value of the *α*−band relative power 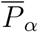 during the plateau duration, we apply the discretized sum approximation

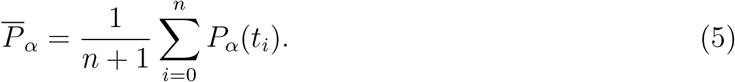
- The *δ*−band mean relative power 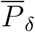 is obtained following the procedure above but taking [0.5, 4] Hz as the reference band.
- The maximum power frequency *f*_*α*_ within the *α*−band is the frequency for which the power is maximal:

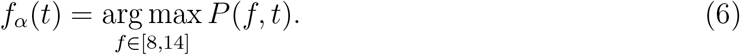

We then approximate the mean *α*_*max*_ by using the discretized sum

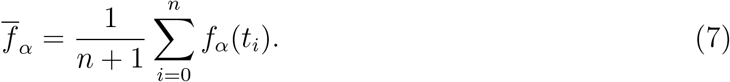

### 2.6 State-Chart representation of EEG activity

To provide an interpretable representation of cortical dynamics during anesthesia, we used a proprietary State-Chart algorithm (SignalMed+, Paris, France) that maps EEG activity onto a discrete set of ordered brain states. The State-Chart is constructed from real-time EEG features derived from a single frontal channel. These features include spectral power distributions (in the *α* and *δ* bands), transient patterns such as *α*-suppressions, iso-electric suppressions (IES), and measures of signal fragmentation [7]. The algorithm integrates these descriptors over time to assign, at each time point, a discrete state value ranging from 0 (deep suppression) to 20 (awake).

Each state corresponds to a characteristic EEG regime, including high-frequency activity in awake conditions, predominance of *α* and *δ* oscillations during stable anesthesia, fragmentation of *α* rhythms, and suppression patterns such as IES and burst–suppression. Transitions between states reflect changes in cortical dynamics rather than predefined drug concentration thresholds.

The State-Chart score is updated continuously and provides a time-resolved representation of brain state evolution. In contrast to scalar indices such as BIS, which summarize EEG activity into a single continuous value, the State-Chart explicitly captures transitions between physiological regimes. The detailed implementation of the algorithm, including feature weighting and decision rules, is proprietary. However, all computations rely on standard EEG signal processing and statistical feature extraction, allowing reproducibility of the underlying physiological interpretation.

### 2.7 Computation of BIS and State-Chart scores

For each EEG recording, BIS values from the clinical monitor and the corresponding State-Chart scores were computed continuously over time. To construct the score distributions (shown in Fig. 4), values were extracted at selected time points corresponding to well-defined anesthetic conditions. Because the pharmacological information differs between propofol and sevoflurane, eligibility criteria were defined separately for each agent. For propofol, scores were collected from 10 min after the first non-zero target concentration, allowing stabilization of drug distribution, and until the target concentration returned to zero.

For each concentration level, we additionally distinguished whether the preceding concentration step was lower (ascending phase, denoted “Up”) or higher (descending phase, denoted “Down”), in order to compare identical concentration values reached through different dosing histories. For sevoflurane, scores were collected exclusively during concentration plateaus, defined as time intervals where the end-tidal concentration remained stable and reliably measured. At each eligible time point, paired BIS and State-Chart scores were recorded. These values were then grouped according to the corresponding anesthetic concentration and used to compute the distributions.

### 2.8 Comparing State-Chart scores (SCS) vs BIS

For propofol, scores were collected from 10 min after the first non-zero target concentration, ensuring stabilization of drug distribution, and until the target concentration returned to zero. For sevoflurane, scores were collected exclusively during concentration plateaus, defined as time intervals where the end-tidal concentration remained stable and accurately measured. To align the state chart scores (SCS) with the 1 Hz sampling frequency of the Bispectral Index (BIS), the state chart time-series, originally sampled every 10 s, was upsampled to 1s using a piecewise constant scheme. State chart values were then separated into the 7 values 30, 35, 40, 45, 50, 55, 60. Finally, the BIS score corresponding to the time points within each state chart scores were grouped into five respective cohorts for analysis. BIS values were then binned into four distinct ranges: [40, 45[, [45, 50[, [50, 55[, and [55, 60[. Finally, the state chart score corresponding to the time points within each BIS interval were grouped into four groups.

#### Statistical permutation procedure to compare sevoflurane vs propofol on EEG spectral signatures

To test whether the hypnotic concentration and EEG parameters differs between propofol and sevoflurane, we compared the concentration-effect curves. The dose concentration was first normalized *c*_norm_ = (*c* − *c*_min_)*/*(*c*_max_ − *c*_min_). For each measured quantities (e.g. relative *α* power, mean *α* suppression proportion,etc..), we fitted a locally weighted regression (`LOESS, frac=0.6, it=0; statsmodels.nonparametric.smoothers lowess.lowess`) to obtain a continuous curve from *c*_norm_. To quantify the difference between the two drugs, we computed the mean squared difference

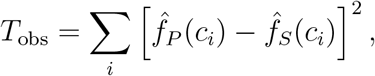

where 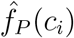 and 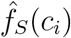 are the interpolated concentration-response curves curves of propofol and sevoflurane respectively at concentrations *c*_*i*_. To evaluate the difference between the groups, we use a permutation test: for each permutation *ω* of *n* = 500 permutations, patients were reassigned to pseudo-drug labels. The mean squared difference *T* ^∗^(*ω*) was recomputed on the relabelled data. The empirical p-value is computed as *p* = (#{*T* ^∗^(*ω*) ≥ *T*_obs_} + 1)*/*(500 + 1).

## 3 Results

### 3.1 EEG signatures at fixed propofol target concentration

We first presented (Fig.1A-B), EEG signatures and spectrograms for propofol and sevoflurane during two independent anesthesia procedures. We further computed the proportion of suppressions of the alpha band (yellow) and iso-electric suppression (red) (Fig.1C-D), that can be compared with concentration of the hypnotic (Fig.1E).

**Figure 1:**
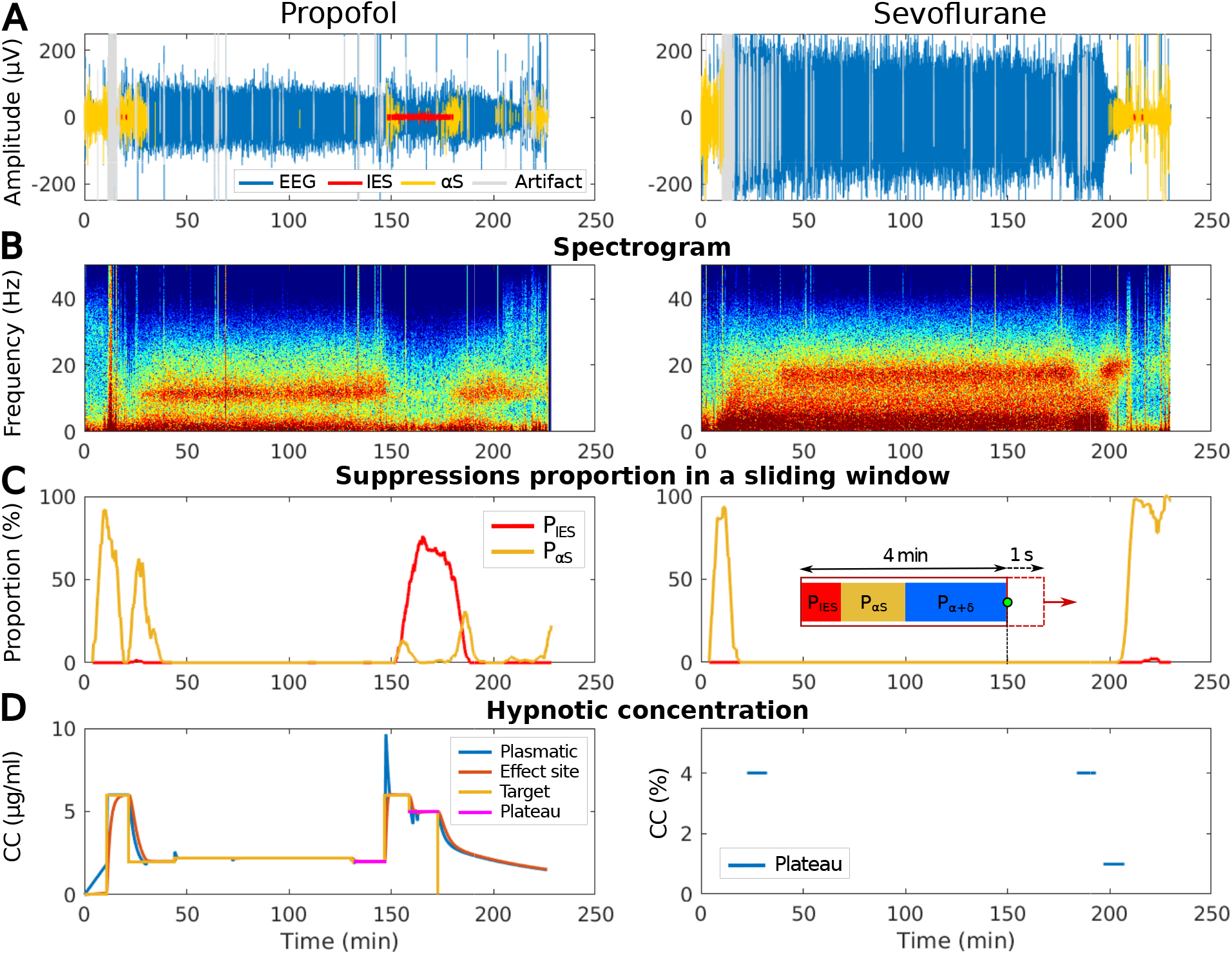
Difference of EEG waveform, spectrogram, suppressions proportion and hypnotic concentrations between propofol and sevoflurane. **(A)** EEG signal (blue) during propofol induced GA segmented in iso-electric suppressions (IES) (red), alpha-suppressions (*α*S) (yellow) and artifacts (grey). **(B)** Spectrogram (time-frequency) representation of the signal shown in (A). **(C)** Proportion of IES (red) and *α*S (yellow) evaluated in a 4 minutes sliding window and 1 second overlap. **(D)** Hypnotic drugs concentrations–propofol (left)–are shown with the following concentration target (yellow) adjusted by hands, the plasma (blue) and the effect site (orange) concentrations are computed according to pharmacokinetics and pharmacodynamics (PK/PD) models and the plateau is shown in violet. For sevoflurane (right), only measured end-tidal concentration plateaus (blue) are shown.

**Figure 2:**
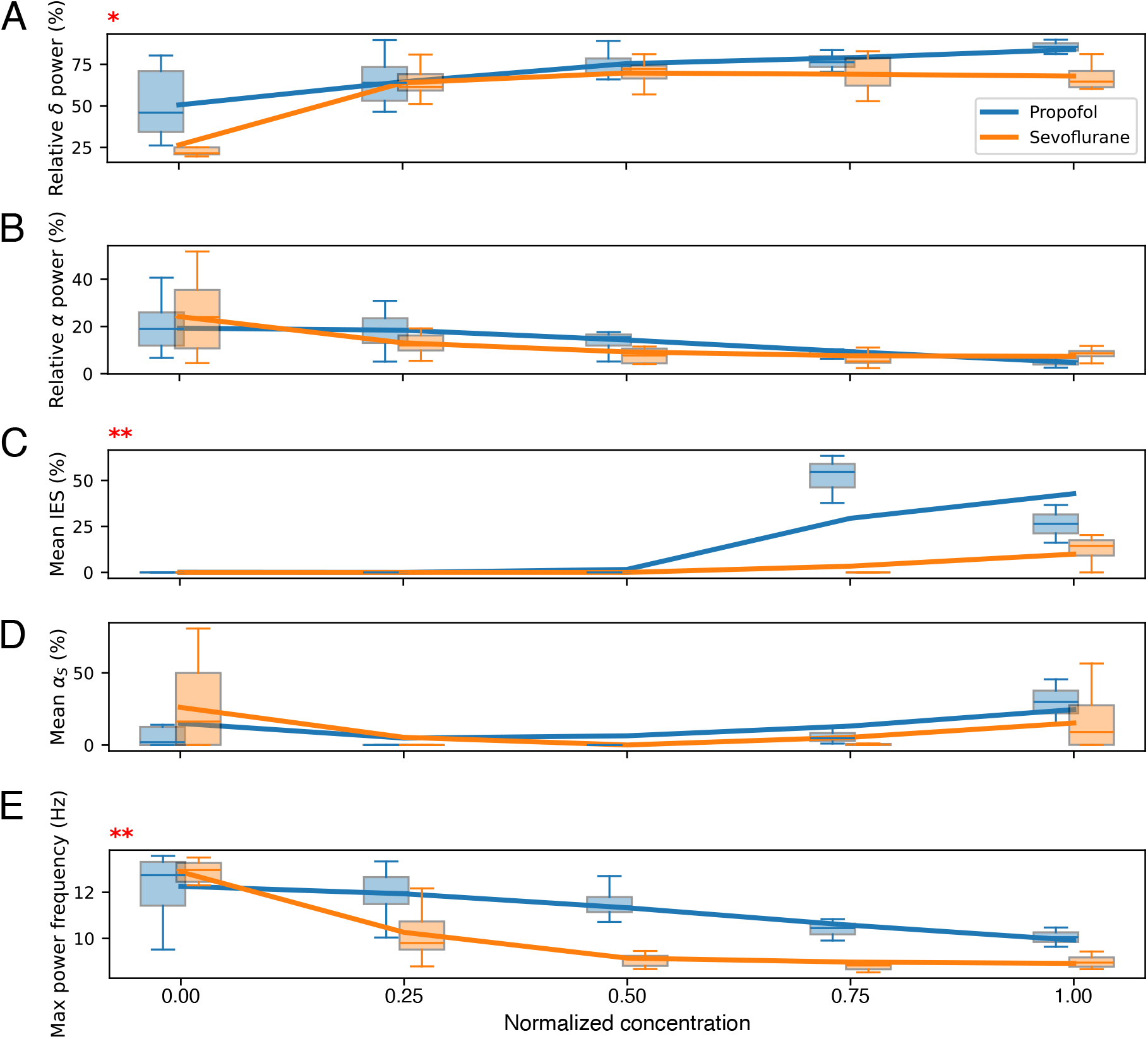
Distribution of IES, *α*S, band power versus propofol target concentration (left) and fraction of expired sevoflurane (right) plateaus. Boxplots (by concentration rank, per drug) with LOESS fits overlaid, for propofol (blue) and sevoflurane (orange). Red asterisks indicate the significance of the LOESS permutation test for that parameter (* *p* < 0.05, ** *p* < 0.01, *** *p* < 0.001); no marker indicates *p* ≥ 0.05. **(A)** Relative *δ*−band power distribution. **(B)** Relative *α*−band power distribution. **(C)** Mean IES proportion. **(D)** Mean proportion *α*S. **(E)** Average maximum power frequency within the *α*−rhythms range.

To further characterize the EEG statistics, we computed the relative *δ*−band power *P*_*δ*_*/P*_*tot*_, where *P*_*tot*_ is the power computed across the frequency range 0.5-30 Hz. We found that the relative *δ*−band power increased along with the concentration (Fig.3.1) for propofol, while it reached a plateau for sevoflurane at an average of 3% end-tidal concentration. In parallel, the relative *α*−band power decreased as the concentration increases, reaching 5% of relative power when the predicted concentration reaches 6 *µg*.*ml*^−1^ (Fig.3.1). In addition, the relative *δ*−power also reached a concentration plateau at around 3% sevoflurane after decreasing. These results show a clear difference between sevoflurane vs propofol.

**Figure 3:**
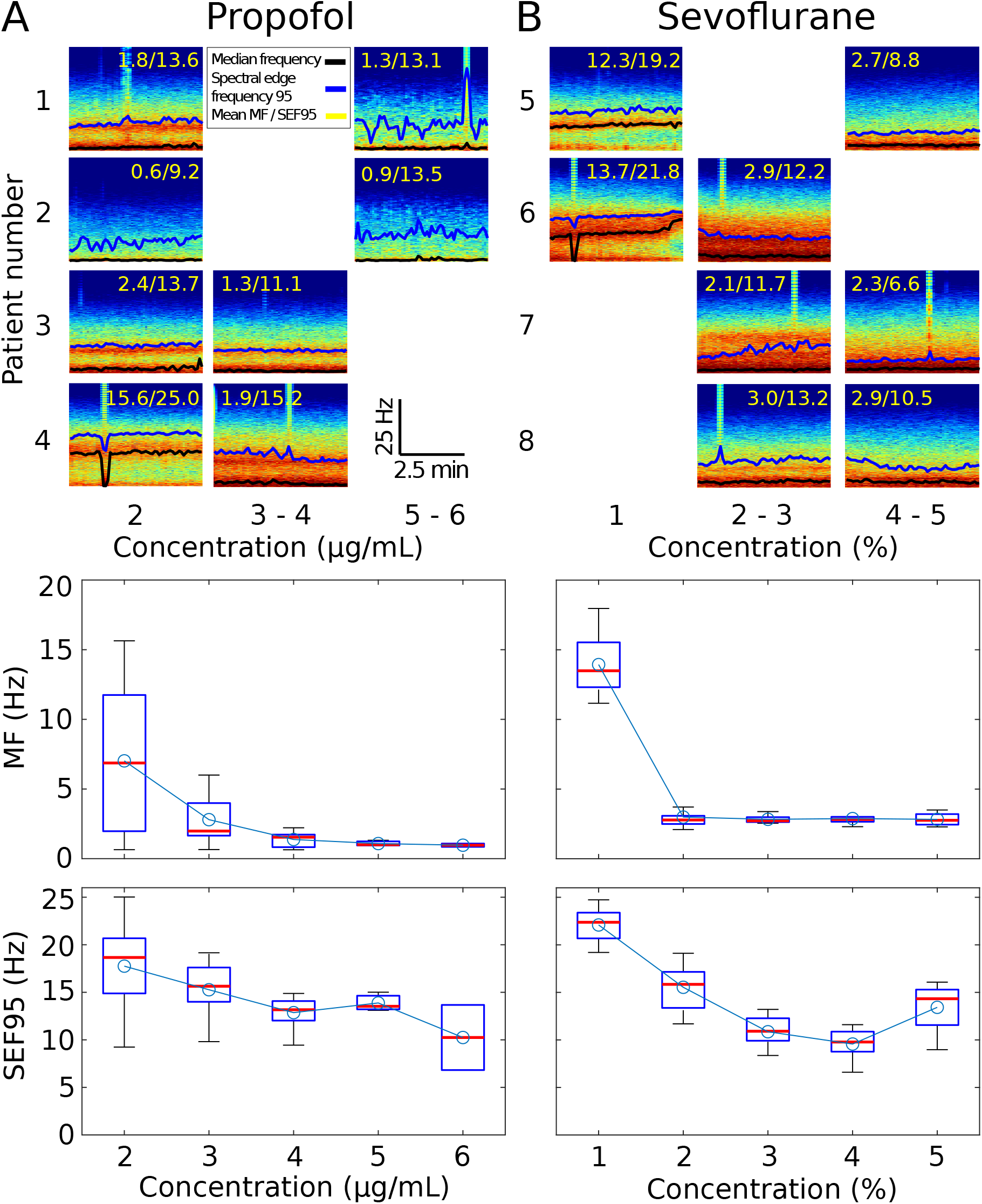
Spectrogram profiles with Median Frequencies (MF) and Spectral Edge Frequency 95 (SEF95) at different plateaus concentration for. **(A)** Propofol, **(B)** Sevoflurane. The Median Frequency (black) and spectral edge frequency 95 (blue) are shown over the spectrogram along with their mean values (yellow). The corresponding boxplots are shown below.

To further explore the arrival of deep sedation with concentration, we decided to detect the presence of iso-electric suppressions (IES) and suppression epochs of the alpha band (*α*S). We report here a sudden increase of IES with both hypnotics. It is however not clear why the mean proportion of IES increases from 5 to 6 *µg*.*ml*^−1^ in the case of propofol. In parallel, we found that the mean proportion of *α*S increased continuously with respect to propofol concentration in the [2-6] *µg*.*ml*^−1^ range. *α*S increased to a maximum after 3% of sevoflurane. At this stage, there were no clear marks of anticipating IES from the dynamics of *αS* (Fig.3.1C). This increase is not necessarily associated with an increase in the mean IES proportion. Instead, IES appeared predominantly for concentration starting at 5 to 6 *µg*.*ml*^−1^. Finally, we observed that the mean of the frequency associated to the maximum power of the *α*−band decreased continuously from 12.3 to 10 Hz (Fig.3.1E) as propofol concentration increases from 2 to 6 *µg/ml*, a situation that is comparable with sevoflurane.

### 3.2 EEG spectral signatures at fixed fraction of hypnotic

To explore the diversity of the spectral signatures of propofol vs sevoflurane, we decided to plot the spectrograms at fixed fraction of hypnotic concentration (Fig.3): While for low concentrations, the spectrogram profile can be different across patients, we found that the SEF95 and median frequency were statistical different for the two hypnotics. Indeed, the median frequency is in average higher for sevoflurane ([13.9, 3.0, 2.8, 2.9, 2.8] Hz) from 1 to 5%) compared to propofol ([7.0, 2.8, 1.4, 1.1, 1.0] Hz from 2 to 6 *µg*.*ml*^−1^) for all concentrations. For concentrations at 3 and 4 units, the SEF95 are in average lower for sevoflurane ([10.9, 9.6] Hz) against propofol ([12.3, 13.9] Hz).

To further evaluate the differences between the concentration-response of propofol and sevoflurane based on the seven EEG spectral signatures (relative delta power (*P*_*δ*_), relative alpha power (*P*_*α*_), mean iso-electric suppression (IES), mean alpha suppression proportion, frequency of maximum alpha-band power, median frequency (MF), and spectral edge frequency 95% (SEF95)), we performed a permutation test (see Method section) on each response. We found that four of the seven EEG parameters (see Table. 2) showed a significant difference between propofol and sevoflurane concentration-effect curves (*p* < 0.05): relative delta power, mean IES, alpha peak frequency, and median frequency, with mean IES and alpha peak frequency reaching the strongest evidence (*p* = 0.006 and *p* = 0.002 respectively). Relative alpha power, Mean alpha suppression proportion, and SEF95 did not reach significance (*p* = 0.188, 0.595, and 0.138), indicating that for these three parameters the concentration-effect relationship could not be distinguished.

**Table 1:** Patients demographics, EEG and statistical features.

| Variables | 89 patients |
| --- | --- |
| Age (mean ± std) | 14.5 ± 7.2 yr |
| Age (range) | 6-38 yr |
| Gender (female/male) | 36 %/64 % |
| Pubertal (pre/post) | 56 %/44 % |
| Concentration plateau (propofol/sevoflurane) | 66/54 |

**Table 2:** LOESS permutation test comparing propofol vs. sevoflurane concentration-effect curves, by EEG parameter.

| EEG parameter | Permutation $p$ |
| --- | --- |
| Relative delta power ($P_\delta$) | 0.014 |
| Relative alpha power ($P_\alpha$) | 0.188 |
| Mean IES | 0.006 |
| Mean αS proportion | 0.595 |
| Alpha peak frequency | 0.002 |
| Median frequency (MF) | 0.010 |
| Spectral edge frequency 95% (SEF95) | 0.138 |

### 3.3 BIS versus State-Chart across propofol concentrations

To compare the conventional BIS index with an interpretable EEG-derived representation, we used a proprietary State-Chart algorithm that projects brain activity onto 21 ordered depth states ranging from deep suppression (state 0) to wakefulness (state 20) (Fig. 4A). Decreasing state values therefore indicate increasing hypnotic depth. Each state is associated with characteristic EEG patterns, including high *γ*/*β* activity in awake conditions, predominance of *α* and *δ* rhythms during balanced anesthesia, fragmentation of the *α* band, and finally isoelectric suppressions (IES), burst–suppression activity, and flat EEG at excessive depth. We first compared the temporal evolution of BIS and State-Chart scores during propofol anesthesia using a sliding window of 60 s with 30 s overlap (Fig. 4B). Although both indices followed the progression from induction to maintenance, the State-Chart score is significantly more stable over time and the distribution of temporal variances was markedly lower for the State-Chart than for BIS (State-Chart: 0.5 ± 0.6; BIS: 4.0 ± 2.9; KS statistic = 0.921, *p* = 9.02 × 10^−90^; Fig. 4C), indicating a more compact representation of the ongoing cortical state. We then analyzed the distributions of BIS and State-Chart scores across propofol concentrations (2– 6 g/ml; Fig. Fig. 4D), where we differentiate whether for the previous state, the concentration was high (UP)or lower (Down) that the present one. Both indices decreased with concentration, consistent with increasing anesthetic depth. However, the State-Chart distributions were narrower and more structured than the corresponding BIS distributions at all concentration levels. In particular, at light-to-moderate propofol concentrations (2–3 g/ml), the State-Chart showed reduced dispersion compared with BIS, suggesting a more reproducible description of stable anesthesia. At higher concentrations (4–6 g/ml), the State-Chart scores shifted toward lower values, corresponding to deeper cortical states associated with *α*-fragmentation, IES, and burst–suppression regimes. Overall, these results indicate that the State-Chart provides a more stable and refined representation of the concentration-effect relationship than BIS during propofol anesthesia. A similar reduction in temporal variance was also observed during sevoflurane anesthesia (Fig. Fig. 4E–G), supporting the robustness of the State-Chart representation across hypnotic agents.

**Figure 4:**
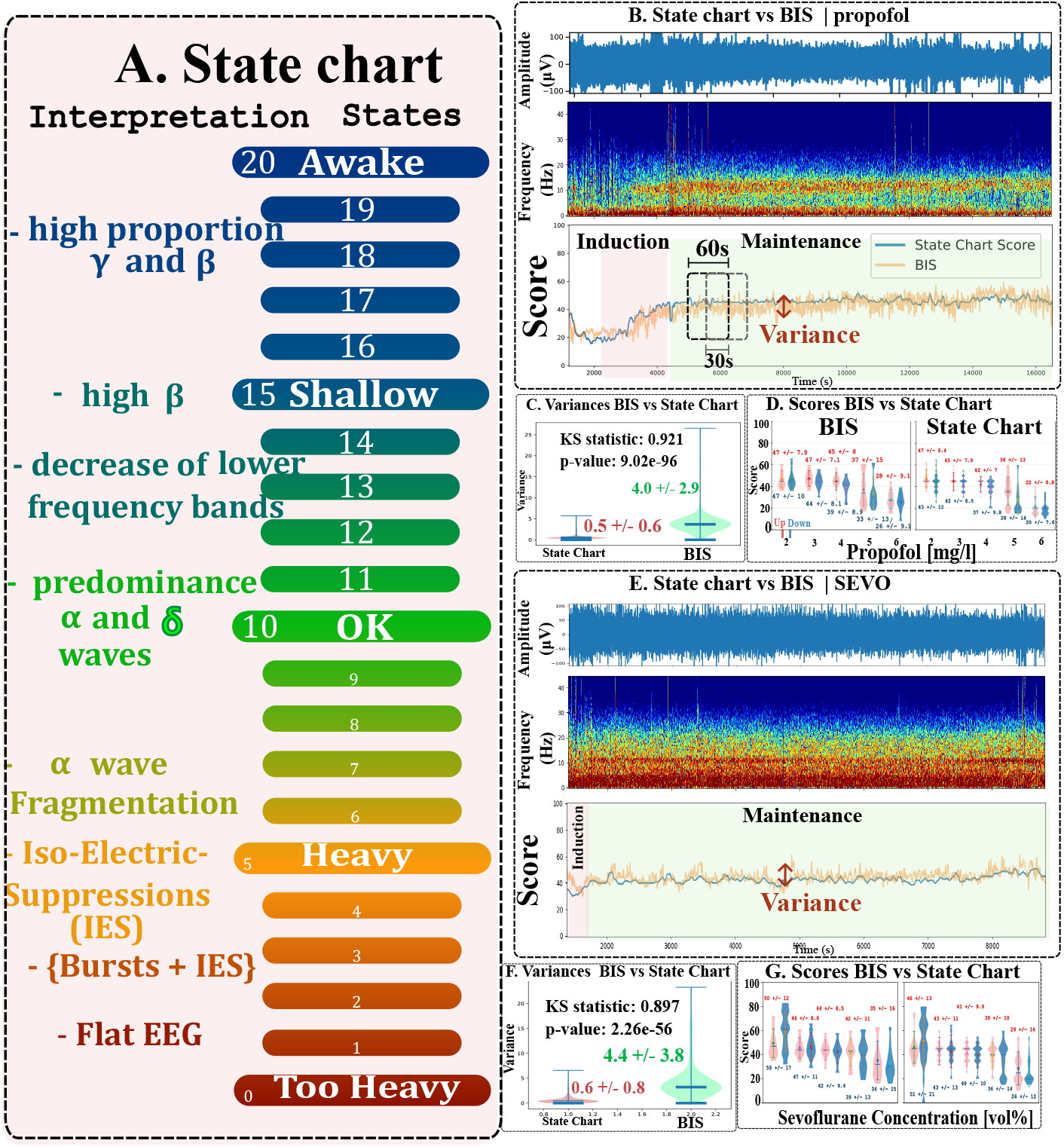
State-Chart representation versus BIS during propofol and sevoflurane anesthesia. **(A)** State-Chart maping EEG activity onto 21 ordered states ranging from wakefulness (state 20) to very deep anesthesia (state 0), with intermediate regimes corresponding to shallow sedation, balanced anesthesia with predominant *α* and *δ* activity, *α*-fragmentation, iso-electric suppression (IES), burst–suppression, and flat EEG. **B.** EEG signal during propofol induced anesthesia (top), spectrogram during induction and maintenance (middle) and BIS vs state chart score over time (bottom). The variances is computed over a sliding windows of 60s with 30 s (50%) overlap. **C**. Distribution of the state chart (left) vs BIS variance (rigth). KS test was performed to quantify the difference in distributions.(0.5 ±0.6 vs 4.0 ± 2.9; KS statistic = 0.921, *p* = 9.02 × 10^−96^) **D**. Distribution of the score for the BIS (left) and state chart (right) at different ranges on effective propofol concentrations for upper (red) vs lower change (blue). Scores are computed on a total of 86 patients, with mean (blue) of the distributions and below the standard deviation (black). **(E)** Propofol recording showing raw EEG, spectrogram, and simultaneous time courses of BIS and State-Chart score during induction and maintenance. **(F)** temporal variance was computed over 60-s sliding windows with 30-s overlap. The State-Chart exhibited markedly lower variance than BIS for sevoflurane (0.6 ± 0.8 vs 4.4 ± 3.8; KS statistic = 0.897, *p* = 2.26 × 10^−56^). **(G)** Distribution of BIS and State-Chart scores across increasing sevoflurane end-tidal concentrations (vol%), showing a progressive decrease in both indices with increasing hypnotic depth, with narrower and more structured score distributions for the State-Chart.

Finally, as the State Chart is decomposed in multiple discreet states, we evaluated the correspondence with BIS. For that purpose, across BIS ranges, we found that a significant proportion of the state chart scores fell below the lower bound of the corresponding BIS interval (Fig. 5). The state-chart often distinguish specific sub-groups showed for BIS intervals of size 5, with a strong concentration of scores around 45, reflecting a preponderance of delta and alpha spectral activity, with delta waves remaining consistently dominant over alpha. Conversely, the reciprocal curve (BIS vs SCS) (Fig. 5) showed a monotonic increase of BIS median values with the unify state chart score. This median is not necessarily centered around the state chart value (scaled by five for direct comparison with BIS).

**Figure 5:**
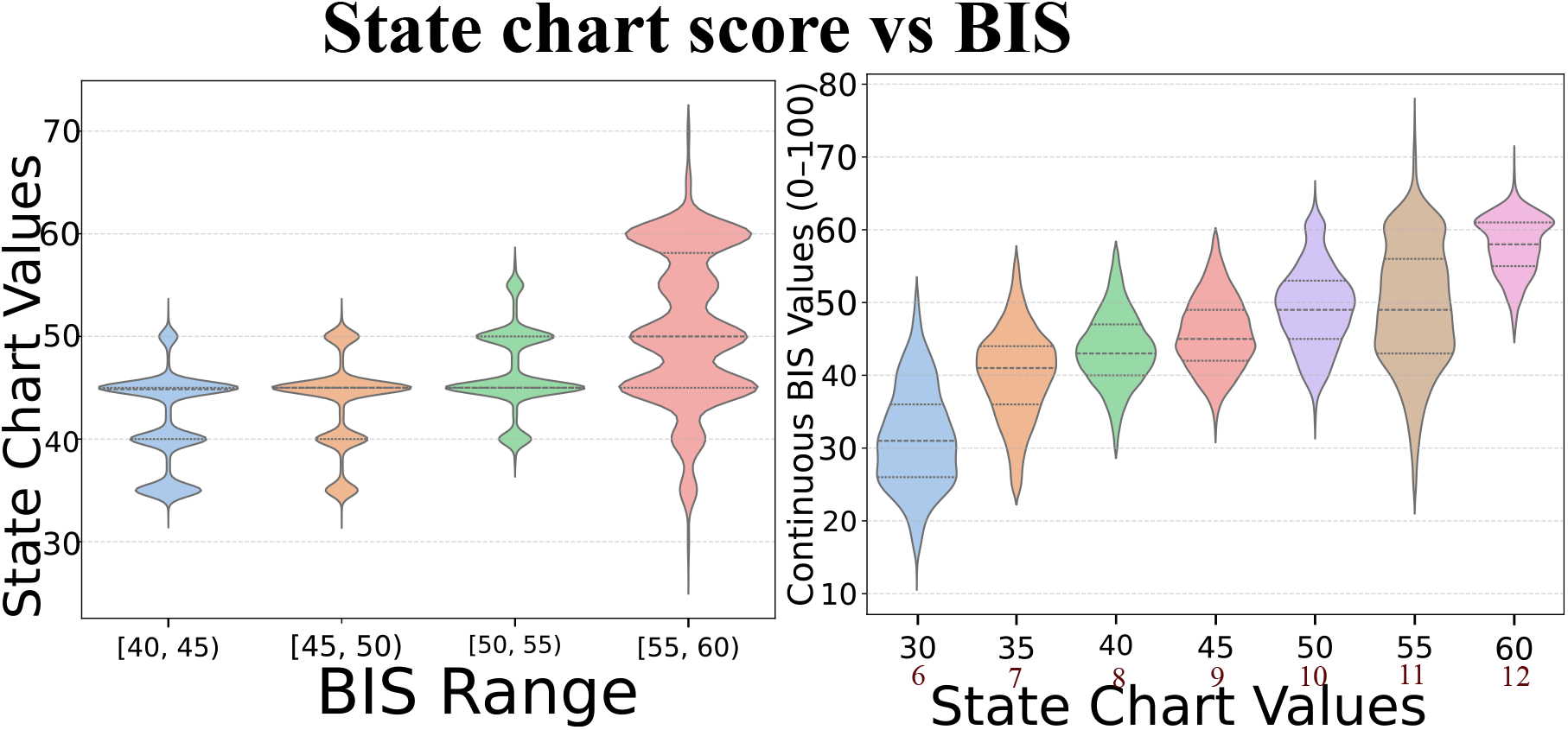
SCS vs BIS and vice versa. Right: from left to right four violin plots corresponding to four non overlapping BIS ranges [40, 45[, [45, 50[, [50, 55[and [55, 60[. that do not overlap. Left: From left to right seven violin plots corresponds to the state-chart values 30, 35, 40, 45, 50, 55, 60 in BIS scaling or 6, 7, 8, 9, 10, 11, 12 in original scaling.

## 4 Discussion and conclusion

In this manuscript, we highlighted the differences between propofol and sevoflurane anesthetic concentrations and presented also the spectral properties of the EEG signal. We computed several parameters, such as the power of the *α*−and *δ*−bands for various sevoflurane and propofol concentrations. To evaluate the depth of anesthesia, we also computed the fraction of time in *α*−suppressions and isoelectric suppressions (Fig.3.1A-B). When propofol concentration increased, the power of the *α*−band decreased, while the *δ* power became dominant (Fig.3.1C-D). However, we did not detect any general trends where the emergence of *α*S would systematically anticipate the appearance of IES. It seems that IES suddenly appear for a specific concentration, suggesting that at a hypnotic concentration high enough, a switch could occur of the dominant brain oscillations. Finally, the spectrograms further reveal a clear variability across patients (Fig.3).

When we compared the propoofol vs sevoflurane effects on the EEG, we found no particular signature differences at low concentration of both hypnotic with a high variability between the spectrogram profiles. At intermediate concentrations ([3-5]*µg*.*ml*^−1^ vs. [2-4]%), the higher median frequency (MF) but lower SEF95 for sevoflurane were associated with wider *δ* and *α* bands compared to propofol, where we have a lower MF but higher SEF95 that we associated with narrow bands and thus less signal amplitude at the corresponding bands frequencies. For high concentrations, the higher MF and SEF95 for sevoflurane were associated with a more dominant *δ*−band, while for propofol the signal amplitude was more suppressed at *δ* frequencies. Finally, the present study complement previous studies [9, 14] on propofol and sevoflurane comparison.

While the BIS provides a single scalar estimate of anesthetic depth that varies with hypnotic concentration but does not explicitly describe underlying brain state dynamics, in contrast, the State-Chart Score (SCS) exhibited markedly lower temporal variability, indicating a more stable representation of cortical activity. This reduced variability suggests that the SCS could provide a lower instantenous variability signal for future closed-loop control strategies. Moreover, as propofol concentration increased, the SCS decreased monotonically and, at higher concentrations (6 *µ*g_•_ml^−1^), separated into two distinct statistics (Fig. 4D). Because the State-Chart incorporates the disappearance of *α* activity (states 6–7), low SCS epochs frequently preceded the onset of iso-electric suppressions, indicating an approaching overdosage state. To conclude, the State-Chart Score provides a stable and physiologically interpretable representation of anesthesia depth and may allow individualized monitoring by identifying transition states preceding cortical suppression.

## Data Availability

All data produced in the present study are available upon reasonable request to the authors

## Abbreviations

EEG: ElectroEncephaloGram
GA: General Anesthesia
TCI: Target Controlled Infusion
*α* S: Alpha-Suppression
IES: Iso-Electric Suppression;

## Acknowledgments

D.H. research is supported by an ANR-23-CE46-0016 AnalysisSpectralEEG and the European Research Council (ERC) under the European Union’s Horizon 2020 research and innovation programme (grant agreement No 882673) and PoC 101212658.

## Data availability

The datasets generated and analysis in the current study are available from the corresponding authors upon reasonable request.

## References

[1] C. Bennett, L. J. Voss, J. P. Barnard, and J. W. Sleigh, “Practical use of the raw electroencephalogram waveform during general anesthesia: the art and science,” Anesthesia & Analgesia, vol. 109, no. 2, pp. 539–550, 2009.

[2] D. Hight, L. J. Voss, P. S. Garcia, and J. Sleigh, “Changes in alpha frequency and power of the electroencephalogram during volatile-based general anesthesia,” Frontiers in systems neuroscience, vol. 11, p. 36, 2017.

[3] P. L. Purdon, A. Sampson, K. J. Pavone, and E. N. Brown, “Clinical electroencephalography for anesthesiologists: part i: background and basic signatures,” Anesthesiology, vol. 123, no. 4, pp. 937–960, 2015.

[4] M. T. Alkire, A. G. Hudetz, and G. Tononi, “Consciousness and anesthesia,” Science, vol. 322, no. 5903, pp. 876–880, 2008.

[5] I. Constant and N. Sabourdin, “The eeg signal: a window on the cortical brain activity,” Pediatric Anesthesia, vol. 22, no. 6, pp. 539–552, 2012.

[6] P. L. Purdon, E. T. Pierce, E. A. Mukamel, M. J. Prerau, J. L. Walsh, K. F. K. Wong, A. F. Salazar-Gomez, P. G. Harrell, A. L. Sampson, A. Cimenser, et al., “Electroencephalogram signatures of loss and recovery of consciousness from propofol,” Proceedings of the National Academy of Sciences, vol. 110, no. 12, pp. E1142–E1151, 2013.

[7] C. Sun and D. Holcman, “Combining transient statistical markers from the eeg signal to predict brain sensitivity to general anesthesia,” Biomedical Signal Processing and Control, vol. 77, p. 103713, 2022.

[8] L. Zonca and D. Holcman, “Emergence and fragmentation of the alpha-band driven by neuronal network dynamics,” PLoS Computational Biology, vol. 17, no. 12, p. e1009639, 2021.

[9] A. Rigouzzo, L. Khoy-Ear, D. Laude, N. Louvet, M.-L. Moutard, N. Sabourdin, and A. Constant, “Eeg profiles during general anesthesia in children: A comparative study between sevoflurane and propofol,” Pediatric Anesthesia, vol. 29, no. 3, pp. 250–257, 2019.

[10] A. R. Absalom and K. P. Mason, Total intravenous anesthesia and target controlled infusions. Springer, 2017.

[11] A. Rigouzzo, L. Girault, N. Louvet, F. Servin, T. De-Smet, V. Piat, R. Seeman, I. Murat, and I. Constant, “The relationship between bispectral index and propofol during target-controlled infusion anesthesia: a comparative study between children and young adults,” Anesthesia & Analgesia, vol. 106, no. 4, pp. 1109–1116, 2008.

[12] M. Dora and D. Holcman, “Adaptive single-channel eeg artifact removal for real-time clinical monitoring,” IEEE Transactions on Neural Systems and Rehabilitation EngineeringF, 2022.

[13] M. Dora, S. Jaffard, and D. Holcman, “The wqn algorithm to adaptively correct artifacts in the eeg signal,” Applied and Computational Harmonic Analysis, vol. 61, pp. 347–356, 2022.

[14] O. Akeju, M. B. Westover, K. J. Pavone, A. L. Sampson, K. E. Hartnack, E. N. Brown, and P. L. Purdon, “Effects of sevoflurane and propofol on frontal electroencephalogram power and coherence,” Anesthesiology, vol. 121, no. 5, pp. 990–998, 2014.

